# Target height influences shoulder and elbow non-use after stroke during reaching with the hand : A brief communication

**DOI:** 10.1101/2022.12.20.22283739

**Authors:** Germain Faity, Denis Mottet, Jérôme Froger

## Abstract

**Background:** During hand reaching movements in people with stroke, the coordination of trunk, shoulder, and elbow muscles changes as a function of target height. However, it is not known whether target height also influences non-use, defined as the difference between two coordinations aiming at the same target.

**Methods:** Twenty-two individuals with stroke (> 1 month) completed the Proximal Arm Non-Use (PANU) test in two conditions: high target (80 cm) and low target (67 cm). Elbow, shoulder, and trunk use was recorded using motion capture.

**Results:** Trunk compensation and non-use of the shoulder and elbow joints were found to depend on target height.

**Conclusions:** Because trunk bending forward goes against the need to elevate the hand, a sufficiently low target is necessary to unmask the presence of shoulder-elbow non-use. We provide novel recommendations for assessing compensations and non-use during hand reaching. Clinical Trial: NCT04747587.

## INTRODUCTION

Many people with stroke may underuse their paretic limb despite sufficient recovery, leading to long-term negative effects (Jones T. A., 2017; Taub et al., 2002). The Proximal Arm Non-Use (PANU) test was designed to measure non-use of the proximal arm during seated reaching tasks, specifically by quantifying the amount of trunk recruitment that could be replaced by increased shoulder-elbow recruitment (Bakhti et al., 2017). Previous research has shown that target height impacts trunk compensations during reaching tasks (Reisman & Scholz, 2006; Valdés et al., 2017), but it is unknown how target height may affect the PANU score.

## METHODS

This study included 20 stroke survivors (11 males, age 61 ± 10 years; 13 right hemisphere strokes, 7 left hemisphere strokes; stroke onset greater than 1 month; able to touch the opposite knee with the paretic arm while sitting). Written informed consent was obtained from all participants before their inclusion. This study was performed in accordance with the 1964 Declaration of Helsinki. The Ethics Committee of Nimes approved the study protocol (N°ID-RCB: 2020-A02695-34, Clinical Trial: NCT04747587).

Participants completed the PANU test at both high (80 cm) and low (67 cm) target heights, with the order of target heights randomized. The distance to the target was adjusted for each participant to match the length of their outstretched active arm, measured from the medial axilla to the distal wrist crease. For each target height, participants first completed a reaching task spontaneously (Spontaneous Arm Use, SAU), followed by a reaching task in which they minimized trunk use to maximize shoulder and elbow use (Maximal Arm Use, MAU) (Bakhti et al., 2017). In the MAU condition, the experimenter lightly touched participants’ shoulders to encourage minimal trunk movement. The PANU score is calculated as the difference in Proximal Arm Use (PAU) between the SAU and MAU conditions:

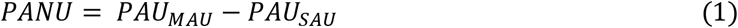

When reach length (Δ_hand_) is *constant between the SAU and MAU conditions*, the PANU score can also be expressed in terms of the contributions by the hand and by the trunk:

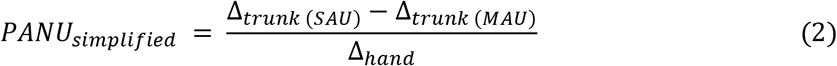

All movements were recorded using Kinect v2, which has been shown to be sufficiently accurate for this type of task (Bakhti et al., 2018).

## RESULTS

A Wilcoxon-Mann-Whitney test found that the PANU score was significantly higher for the low target (12.04 % vs. 5.92 %, U = 235, p = .001) (Fig. 1, panel A). A two-way ANOVA revealed that trunk compensation decreased for maximal arm use (F(1, 106) = 7.464, p = .007) and increased with target height (F(1, 106) = 11.498, p < .001) with no interaction (F(3, 106) = 0.116, p = .735) (Fig. 1, panel B).

**Figure 1.**
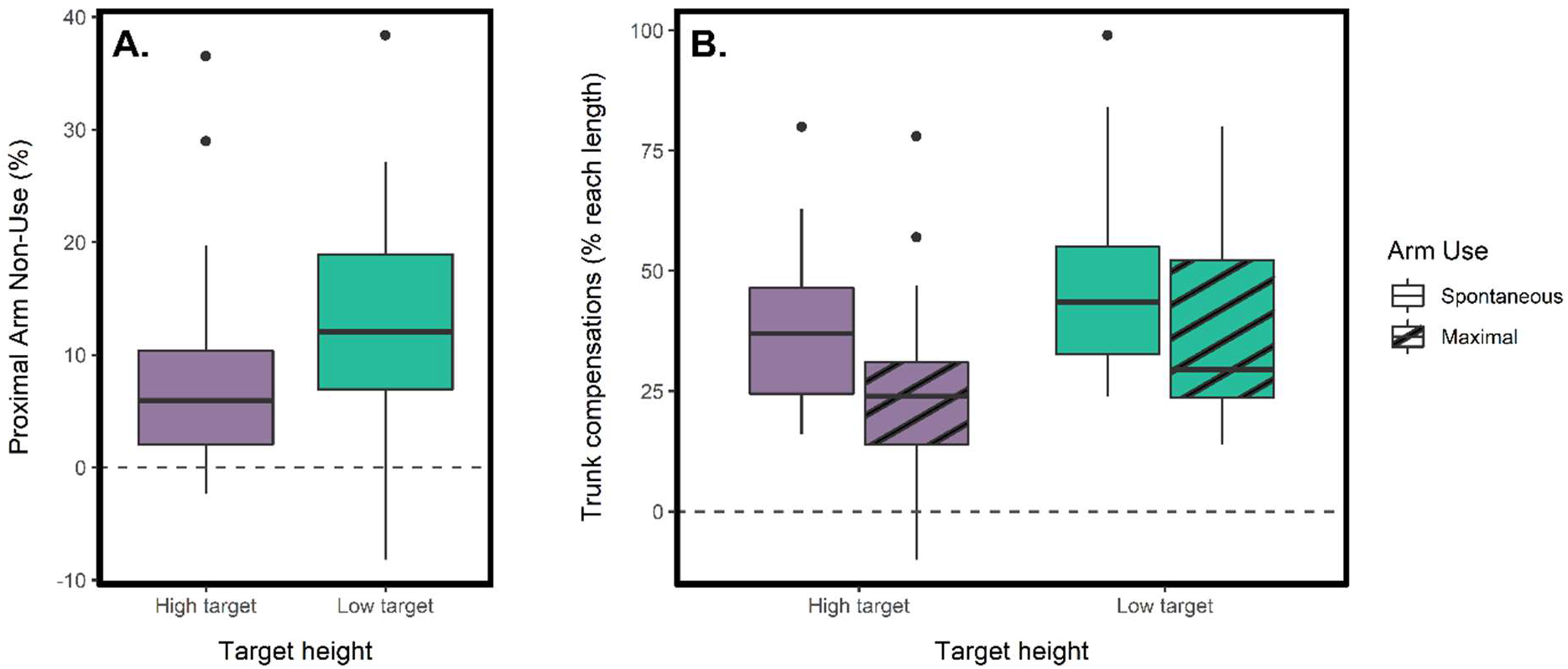
Panel A shows PANU scores for high and low target height conditions, with a lower PANU score observed for the higher target. Panel B illustrates trunk compensations for both arm use and target height conditions, with participants demonstrating a lower level of trunk recruitment for the high target and in the maximal arm use condition.

## DISCUSSION

As previously discussed by Valdés et al. (2017), when the target is low (67 cm), anterior flexion of the trunk allows the shoulder and hand to be advanced close to the target while keeping the elbow flexed and with minimal shoulder flexion (Fig. 2). In contrast, when the target is higher (80 cm), the strategy of trunk bending goes against the need to elevate the hand, requiring some flexion or abduction of the shoulder to reach the target. This results in trunk compensation becoming superfluous in both spontaneous and maximal arm use conditions (i.e., maximal arm use is mandatory in both conditions), leading to a lower PANU score (eq. 1). Conversely, a low target allows patients to spontaneously favour trunk recruitment, which can reveal the presence of shoulder-elbow non-use that is quantified by a higher PANU score.

**Figure 2.**
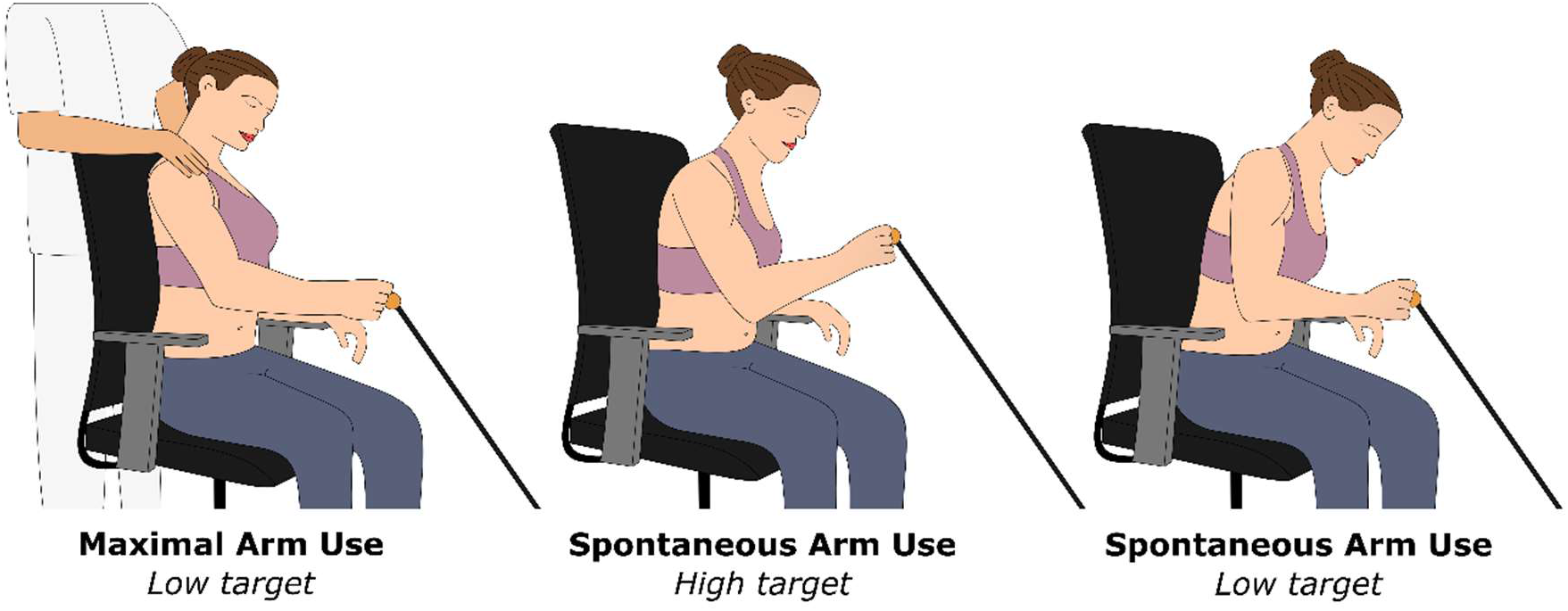
Depiction of three of the four experimental conditions. In the spontaneous conditions, participants were asked to reach the target naturally. In the maximal arm use condition, participants were instructed to minimize trunk use to maximize shoulder-elbow use. Results demonstrate that trunk compensations are reduced in the high target condition compared to the low target condition.

It is important to note that the distance travelled by the hand during reaching is also involved in the calculation of the PANU score (eq. 2). Therefore, if the starting position of the hand is closer to the target and the recruitment of the trunk remains the same, the hand will travel a shorter distance (i.e., a lower Δ_hand_ in eq. 2), which will artificially increase the PANU score. This issue can be particularly relevant for patients who perform the PANU test using their own wheelchair. As the length of the armrests on wheelchairs can vary significantly, it is more appropriate to define the starting position of the hand relative to the participant’s body rather than relative to the armrest of the chair.

Considering the influence of target height and hand starting position, we make the following recommendations when assessing shoulder and elbow non-use with the PANU score:

1. The target should be approximately 80 cm high if the patient is in a wheelchair and 67 cm high if the patient is sitting in a standard chair. This is based on the princeps study (Bakhti et al., 2017) which established PANU standards for the stroke and non-stroke population using a 22 cm height difference between the target and the chair seat.
2. The patient should start the movement with the trunk straight (trunk axis orthogonal to the ground plane) and the distal wrist crease approximately at the level of the xiphoid process on the anteroposterior axis.
3. The patient must reach the target with their hand. Any failed attempts (where the hand does not reach the expected final position) should not be considered.

When these conditions are met, the PANU score can be used to effectively monitor shoulder and elbow non-use, enabling the adaptation of the rehabilitation strategy based on the patient’s recovery.

## Data Availability

All data produced in the present study are available upon reasonable request to the authors.

## ACKNOWLEDGMENTS

This study was supported by Nîmes University Hospital (N°ID-RCB: 2020-A02695-34) and the LabEx NUMEV (ANR-10-LABX-0020) within the I-SITE MUSE.

## DATA AVAILABILITY

The data that support the findings of this study are available upon reasonable request.

